# The early scientific literature response to the novel Coronavirus outbreak: who published what?

**DOI:** 10.1101/2020.03.25.20043315

**Authors:** Davide Gori, Erik Boetto, Maria Pia Fantini

## Abstract

**Introduction:** Recent events highlight how emerging and re-emerging pathogens are becoming global challenges for public health. In December 2019, a novel coronavirus has emerged. This has suddenly turned out into global health concern.

**Objectives:** Aim of this research is to focus on the bibliometric aspects in order to measure what is published in the first 30-days of a global epidemic outbreak

**Methods:** We searched PubMed database in order to find all relevant studies in the first 30-days from the first publication.

**Results:** From the initial 442 identified articles, 234 were read in-extenso. The majority of papers come from China, UK and USA. 63.7% of the papers were commentaries, editorials and reported data and only 17.5% of the sources used data directly collected on the field. Topics mainly addressed were “epidemiology”, “preparedness” and “generic discussion”. NNR showed a reduction for both the objectives assessed from January to February.

**Conclusions:** “Diagnosis” and effective preventive and therapeutic measures were the fields in which more research is still needed. The vast majority of scientific literature in the first 30-days of an epidemic outbreak is based on reported data rather than primary data. Nevertheless, the scientific statements and public health decisions rely on these data.

**Strengths of our study:** This is the first bibliometric research in Pubmed Database on the first 30 days of publications regarding the novel Coronavirus (SARS-nCoV-2) outbreak of 2019.

The vast majority of publication in the first 30-days of an epidemic outbreak are reported data or comments, and only a small fraction of the papers have directly collected data.

**Limitations of our study:** Our research is only PubMed based. It ill be auspicable to consult more than one relevant database in future papers.

In addition, we excluded non-English publications leading to a potential bias due to the fact that the outbreak started in China.

## Introduction

Recent events highlight how emerging and re-emerging pathogens are actually becoming global challenges for public health. [1]

Coronaviruses are enveloped RNA viruses which are broadly distributed either in humans or in a vast majority of other mammals and birds. These viruses have the possibility to cause respiratory, enteric, hepatic, and neurologic diseases. [2, 3] Coronaviruses are highly prevalent in many species. They have a large genetic diversity. Given their RNA, they are susceptible to frequent recombination and mutations of their genomes. In context in which there are increasing human–animal interactions, novel coronaviruses are very likely to emerge periodically. If the newly created cross-species pathogens acquires the ability to infect humans or to be transmitted human to human it can lead to occasional spillover events and epidemics. [4, 5, 6] In the past years two other strains of Coronavirus - severe acute respiratory syndrome coronavirus (SARS-CoV) and Middle East respiratory syndrome coronavirus (MERS-CoV) - have emerged as potential public health worldwide threats. [5] SARS-CoV was the causal agent of the severe acute respiratory syndrome outbreaks in 2002 and 2003 in Guangdong Province, China; [7, 8, 9] MERS-CoV was the pathogen responsible for severe respiratory disease outbreaks in 2012 in the Middle East. [10]

On 31 December 2019, the World Health Organization (WHO) China Country Office was informed of cases of pneumonia of unknown etiology detected in Wuhan City, located in the Hubei Province, China, associated with exposures in a seafood and wet wholesale market in the same city. A new type of coronavirus was isolated on 7 January 2020. [11] On 30 January 2020 WHO declared the outbreak to be a Public Health Emergency of International Concern (PHEIC). [12] This novel coronavirus suddenly turned out to be a global health concern for a disease, called Coronavirus disease 2019 (COVID-19), [13] which was characterized as a pandemic by WHO on 11 March 2020. [14]

Starting from the second-half of January 2020, scientific literature has been particularly focused on the description of this new viral outbreak; the main topics addressed were the epidemiological, clinical and virological aspects as well as the possible public health choices necessary to contain the spread of the disease. Nevertheless, several aspects are still unclear and have not been thoroughly explored, leaving grey areas in our knowledge of the disease and of the outbreak.

The aim of this paper is to perform a bibliometric analysis on the first papers published in the early stages of the SARS-CoV-2 outbreak, in order to give a glimpse to the researchers of “who published what” at the very beginning of this Public Health Emergency of International Concern.

## Methods

### Electronic searches

We searched MEDLINE (PubMed) electronic database in order to find all relevant studies. After a preliminary search starting from 1 December 2019, we found out that the first article meeting our inclusion criteria was published on 14 January 2020. [15] [Supplementary materials, Table 1, ID: 1] We subsequently extended our research to the first 30 days after this milestone article, up to 13 January 2020. We decided to scan the reference lists of the reviews retrieved in order to find potential additional pertinent articles, and to test string sensitivity.

The search string used was: *coronavirus* OR Pneumonia of Unknown Etiology OR COVID-19 OR nCoV*. The virus name was updated from “2019-nCoV” to “SARS-CoV-2” by the International Committee on Taxonomy of Viruses on 11 February 2020, [16] yet we performed the search using the term “nCoV” since we presumed that no paper published between 11 February 2020 and 13 February 2020 would have used the term “SARS-COV-2”. To achieve the highest sensitivity, we decided to use only a combination of keywords avoiding Mesh terms. Asterisks are used to truncate words, so that every ending after the asterisks was searched. We placed a language restriction for English, without other limits to the search.

### Data selection and extraction

The papers were than classified on the basis of a standardized list of key information including: Digital Object Identifier number (DOI), title, journal name, day of first publication (online or paper form), country of the first author affiliation, article type, source of the data and topics addressed. The topics addressed were identified according to 8 categories: generic discussion, epidemiology, virology, pathology and clinical presentation, diagnosis, therapy, transmission, preparedness. Articles that addressed more than a topic in an extensive way were accounted for more than one of the 8 topic categories; the papers that could not be classified in any of the 8 chosen topic categories were classified in the “Other” topic category.

Eight of us (FS, EA, EB, EB, FE, FM, FS, SS) independently screened all identified articles. Procedure was carried out through title and abstract scanning in order to verify the inclusion criteria. Any disagreements were resolved through discussion and consensus between the reviewers. If disagreement persisted, another reviewer (DG) was called as tie-breaker.

### Statistical analysis

The “Number Needed to Read” (NNR) is defined as the ratio of the number of retrieved abstracts to the number of the ones which are pertinent for the research purpose. [17] We calculated the NNR to identify the proportion of papers which were not editorials and commentaries, as well as the proportion of papers with directly collected data.

Chi2 test was used in order to test for significant differences in the comparison of percentages, using the statistical software STATA 15 (StataCorp. 2017. Stata Statistical Software: Release 15. College Station, TX: StataCorp LLC).

### Patient and Public Involvement

Patients or the public WERE NOT involved in the design, or conduct, or reporting, or dissemination plans of our research.

## Results

### Affiliations and publication type

A total of 442 papers were retrieved; 234 articles were found to be pertinent (see Figure 1) and read *in extenso* in order to define their main characteristics.

**Figure 1.**
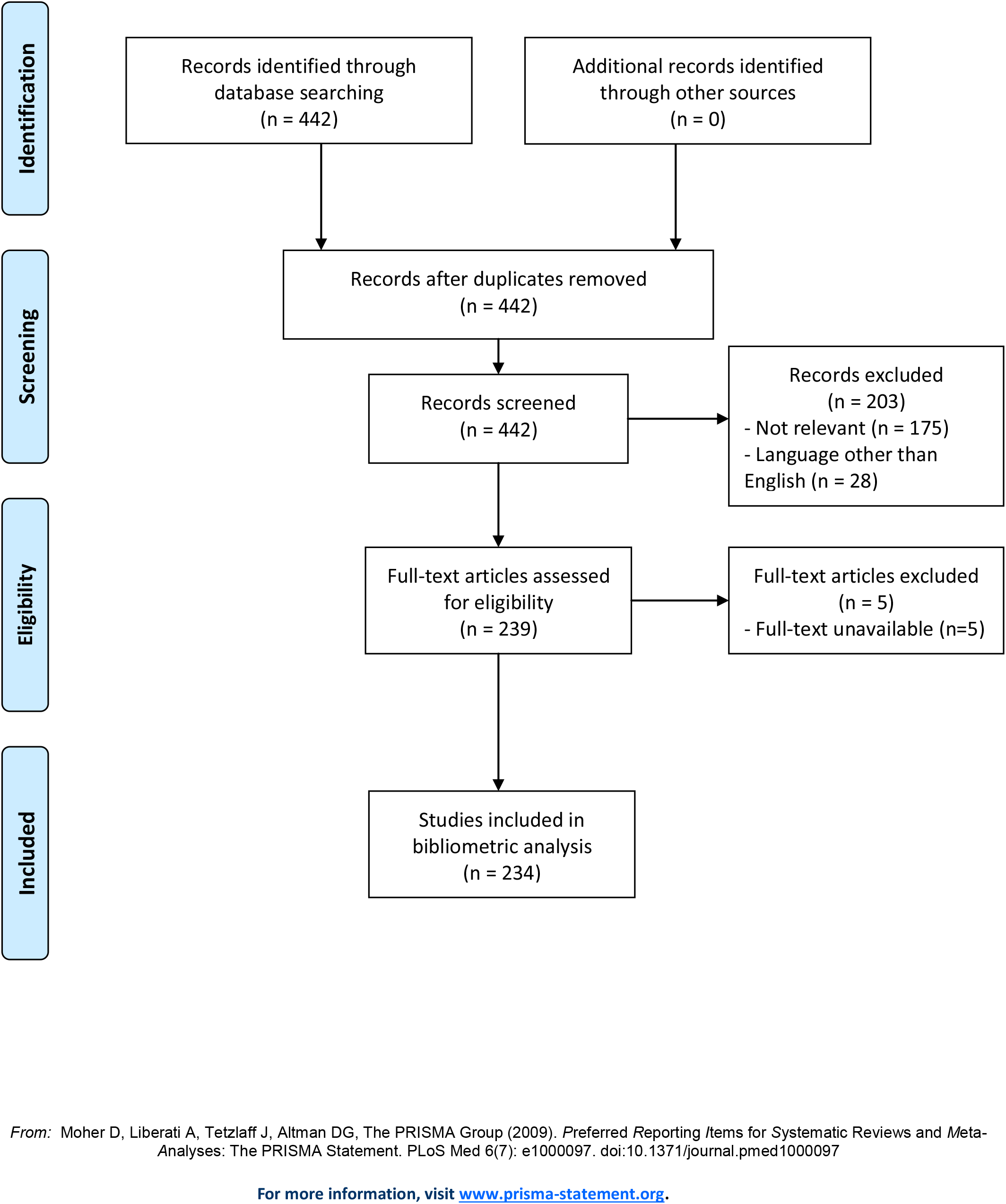
Bibliometric Analysis PRISMA Flow Diagram. **Text:** This PRISMA Flow Diagram shows the process we performed to select the papers included in the bibliometric analysis. Supplementary materials, Table 1 **Title:** Reference, general characteristics and description of the 234 papers selected. **Description:** this table gathers the Identification Number (ID) we assigned to each of the 234 papers, along their distinctive DOI and general information such, as First Author, Journal and Publication Date. Each papers was classified according to the source of the data, the study type, and the topic addressed. **LEGEND** **ID:** Identification Number **DOI:** Digital Object Identifier **GEN DISC:** General Discussion **EPID:** Epidemiology **VIR:** Virology **PATH & CLIN:** Pathology and Clinical Presentation **DIAG:** Diagnosis **THER:** Therapy **TRAN:** Transmission **PREP:** Preparedness **Notes:** - Regarding “Journal Publication Date”, some dates may be consecutive to 13 February 2020; this is due to the fact that some paper were already indexed in MEDLINE database (PubMed) at the moment we performed our search.

The affiliation of the first authors was distributed among 36 countries: 36.3% of the papers (85 out of 234) had a first author with a Chinese affiliation, 18.4% and 13.7% (43 out of 234 and 32 out of 234) had a first author with an affiliation from UK and USA respectively, while first authors affiliation from Canada accounted for 3.8% of the total (9 out of 234), from Italy for 3% (7 out of 234), and from Australia and Korea both accounted for 2.1% (5 out of 234 each). All the other countries accounted for less than 2% of the first author affiliations (less than 5 out of 234).

63.7% of the papers (149 out of 234) were editorials, commentaries or letters (mainly reported data). 10.7% of the papers (25 out of 234) were secondary papers, mainly narrative reviews, which collected the knowledge available up to that point on some specific topics (i.e. genomics of the virus, transmissibility, etc.). The remaining 25.6% (60 out of 234) were original primary studies: among these, case reports accounted for 43.3% of the total, while in vitro or in vivo studies or genomic studies accounted for 21.7% of the total. The remaining primary studies were cohort studies, case control studies and surveys.

Chinese first authors published more original data and primary studies than authors from the UK and the USA (32.9% vs 4.7% of the UK and 6.3% of the USA; p<0.001), who published mostly editorials and commentaries.

Of note, 61.5% of the analysed papers used reported/non original data and 15.8% used official data. Conversely, only 17.5% of the papers used data which were directly collected on the field. In 5.2% of the papers the source of the data was not clearly specified.

The supplementary materials (Table 1), available online, report all the findings of this review with the reference and description of the 234 papers selected for the bibliometric analysis.

### Topics and content of the papers published

“Generic discussion” papers were the most frequent ones (29.5% of the total), and consisted mostly of editorials and commentaries, with no original data. The second most frequent topic was “Preparedness” (23.1%): 83.3% of these papers consisted of commentaries, and only 1 paper was a primary study. The “Epidemiology” topic was addressed in 15.4% of the papers, and only 2 (5.6%) were primary studies. The lack of primary studies could be observed also for the “Virology” topic (14.1%) and the “Transmission” topic (12.8%), with only 12 (36.4%) and 6 (20%) papers reporting original data, respectively. On the other hand, the “Pathology and clinical characteristics” topic (12.8%) consisted mostly of primary studies (22, 73.3%). “Therapy” topic was addressed in 12% of the papers, with 9 papers (33.3%) being primary studies. The least addressed topic was “Diagnosis” (9.4%), with 12 papers (54.5%) reporting original data. Finally, 3% of the papers were classified in the “Other” topic category.

As shown in supplementary materials, the 22 original papers which addressed the clinical characteristics of patients with COVID-19 [Supplementary materials, Table 1, IDs: 21, 22, 54, 68, 69, 79, 128, 135, 148, 150, 152, 153, 154, 165, 173, 175, 182, 188, 196, 208, 225, 229] have been primarily published in the second half of the time frame selected and were mainly case reports; the majority of the first authors had a Chinese affiliation.

The 12 papers which reported original data on the diagnosis topic [Supplementary materials, Table 1, IDs: 22, 54, 68, 69, 97, 128, 148, 165, 198, 199, 206, 218] addressed CT scan imaging features of COVID-19 patients from case reports/series. An Eurosurveillance communication report [18] [Supplementary materials, Table 1, ID: 218] assessed the required expertise and capacity for molecular detection in specialised laboratories into the European Union/European Economic Area countries. This paper emphasized the need for countries to put in place strong measures in order to detect and laboratory-confirm cases early. It was also highlighted the need to perform molecular testing (RT-PCR, as also indicated by the WHO [19]) on different specimens including: nasopharyngeal swabs, bronchoalveolar lavage, oropharyngeal swab, nasopharyngeal aspirate, sputum, (endo) tracheal aspirate and nasal wash. The communication report also underlined the need for a clinical validation of the test specificity and sensitivity. If nasopharyngeal and oropharyngeal swabs are the recommended types of specimen for diagnostic testing an interesting brief report published by Chinese authors [20] [Supplementary materials Table ID: 206] showed the opportunity of using saliva specimens for diagnosis, monitoring, and infection control in patients with SARS-CoV-2 infection.

Concerning the “Therapy” topic, 9 original studies [Supplementary materials, Table 1, IDs: 68, 91, 135, 148, 152, 165, 167, 197, 222] were published in the second half of the time frame selected, mostly by authors with a Chinese affiliation. These papers addressed several aspects of COVID-19 treatment, such as interferon inhalation, use of and efficacy of antiviral drugs, potential repurposing treatments with angiotensin receptor blockers, use of antibiotics for bacterial co-infections, and respiratory support therapy (e.g. oxygen saturation, CPAP, invasive mechanical ventilation). Interestingly, one paper published on 4 February 2020 [21] [Supplementary materials, Table 1, ID: 91] investigated the possibility that the receptor that SARS-CoV-2 uses to infect lung cells might be ACE2, a cell-surface protein on lung AT2 alveolar epithelial cells; one of the known regulators of endocytosis for the AT2 cells is the AP2- associated protein kinase 1 (AAK1). The authors suggested the possibility to use high-affinity AAK1-binding drugs to inhibit endocytosis of AT2 cells, such as baricitinib (an oral, targeted synthetic disease-modifying antirheumatic drug used for the treatment of rheumatoid arthritis), in order to reduce both the viral entry and the inflammation in patients.

### Assessment of the NNR

From the second half of January to the first half of February the proportion of editorials and commentaries changed the NNR from 3.6 to 2.6, meaning that we passed from a mean 3.6 papers to read in order to find an non-editorial/commentary to 2.6 papers. The mean chance to read papers with directly collected data increased, with the NNR passing from 6.5 in January to 5.4 in February.

## Discussion

The novel coronavirus infection that emerged two months ago (as already discussed, the new coronavirus was isolated on 7 January 2020) places emphasis on the importance of ensuring that health professionals, researchers and the public have the best possible scientific information.

Bibliometrics is a scientific method widely used in many fields for quantifying and analysing published information. [22] This kind of literature analysis in the first month after the first publication in Pubmed shows a great number of papers published as expected, mainly by authors with a Chinese affiliation, followed by authors with an UK or USA affiliation. The reason for this distribution is ascribed to the fact that China is the country where the virus was originally isolated and the first one with a COVID-19 outbreak. Moreover, we have to point out that, as shown in a commentary published in Nature, in recent years China produced more science PhD than any other country in the world and the Chinese government has been assuming a leading role in supporting its own scientific research. [23] Therefore the research and publication response to the outbreak has been prompt and lively, giving to the scientific community many elements for understanding the new infection. The second country with the highest number of publications was the UK, followed by the USA, probably due to the fact they are the most prolific countries for scientific research. Authors from the UK and the USA wrote mostly editorials and commentaries.

The publication policies of many of the most important scientific journals were adapted to quickly review and publish scientific papers on SARS-CoV-2 and COVID-19. This strategy facilitated the spread of new knowledge on the new infection and on the outbreak. Hence, it is important to have scientific shared papers in order to reduce misinformation which is itself a public health threat. [24]

In the early scientific literature we analysed there is a lack of primary studies with original data, as expected, given the very short period of time considered after the first pertinent publication on the new infection. Nevertheless, editorials, news and commentaries are important to share opinions and the viewpoint of an expert or a panel of experts, rather than on producing new scientific evidence.

Considering the primary studies, as we can see from the results of our review, they mainly addressed clinical characteristics, signs and symptoms of the new infection, giving a glimpse of the variety and severity of the presentation of COVID-19. Further publication and reviews on this topic are needed to summarize the knowledge arising from all the case reports.

Diagnostic tools, except for CT scan, were poorly represented and discussed, especially in terms of molecular diagnosis and standardization of the techniques.

The “Therapy” topic was also poorly represented, since more time is needed for clinical intervention studies.

Analyzing the selected time frame, the percentage of the editorials, news and commentaries has decreased. This was highlighted particularly by our analysis, which shows that from the second half of January to the first half of February the proportion of editorials and commentaries lowered the NNR, thus indicating an increase in the level of evidence produced as well as the increase of primary data availability.

There are several limitations in our study. First, we based our research only on PubMed, that does not index all of the scientific impacted journals. It is strongly recommended to consult more than one relevant database when performing systematic reviews of the literature, [25] even though it has been reported that most of the high-quality articles, like those included in Cochrane Reviews, are indexed in PubMed. [26] In addition, we excluded all the publications not in English. We acknowledge that, given the outbreak started in China, this may have led to a partial selection bias. Nevertheless, papers excluded which were written in Chinese and had an English abstract, were for the greatest part reports of the Chinese Government (data not shown). We considered only the articles published until 13 February 2020, and taking into account that literature is now changing day-by-day for this topic, extending the bibliometric research for the next months would be of paramount importance.

In conclusion, as far as we know, our very early review is the first bibliometric study analyzing the early scientific research output about COVID-19 from a quantitative and descriptive standpoint. Due to the huge amount of interest and concern related to this outbreak, our analysis shows that the scientific publication has been very reactive but still preliminar, with an expected deficit of original data and an excess of editorials and commentaries. Further research is needed to review and synthesize the growing knowledge on SARS-CoV-2 and COVID-19, with a special aim on the most important knowledge gaps in terms of diagnosis and effective preventive and therapeutic measures.

## Data Availability

The authors declare that all the data relevant to the study are included in the article or uploaded
as supplementary information in Supplementary Table 1

## Contributorship statement

D.G. and M.P.F. contributed to the design and implementation of the research. F. S., E.A., E.B., E.B., F.E., F.M., F.S. and S.S. contributed to the data browsing, data extraction and analysis. D.G. helped in the revision of the dubious cases. E.B. designed the graphs and the table. D.G. provided the drafting of the manuscript. E.B. and M.P.F. provided the final revision of the manuscript.

## Data sharing statement

The authors declare that all the data relevant to the study are included in the article or uploaded as supplementary information in Supplementary Table 1

